# Breakthrough Covid-19 cases despite tixagevimab and cilgavimab (Evusheld™) prophylaxis in kidney transplant recipients

**DOI:** 10.1101/2022.03.19.22272575

**Authors:** Ilies Benotmane, Aurélie Velay, Gabriela Gautier Vargas, Jérôme Olagne, Samira Fafi-Kremer, Olivier Thaunat, Sophie Caillard

## Abstract

While the combination of casirivimab-imdevimab (Ronapreve™ Roche Regeneron) has been shown to confer satisfactory protection against the delta variant kidney transplant recipients (KTRs) with COVID-19, it has limited neutralizing activity against the current variants of concern (Omicron BA.1, BA.1.1 and BA.2). In contrast, cilgavimab-tixagevimab combination (Evusheld™, Astra Zeneca) retains a partial neutralizing activity against omicron in vitro. We examined whether preexposure prophylaxis with Evusheld™ can effectively protect kidney transplant recipients (KTRs) against the Omicron variant.

Of the 416 KTRs who received intramuscular prophylactic injections of Evusheld™ (150 mg tixagevimab and 150 mg cilgavimab), 39 (9.4%) developed COVID-19. With the exception of one patient, all KTRs were symptomatic. Hospitalization and admission to an intensive care unit were required for 14 (35.9%) and three patients, respectively. Two KTRs died of COVID-19-related acute respiratory distress syndrome. SARS-CoV-2 sequencing was carried out in 15 cases (BA.1, n = 5; BA.1.1, n = 9; BA.2, n=1). Viral neutralizing activity of the serum against BA.1 variant was negative in the 12 tested patients, suggesting that this prophylaxis strategy provides insufficient protection against this variant of concern.

Preexposure prophylaxis with Evusheld™ does not adequately protect KTRs against Omicron. Further clarification of the optimal dosing can assist in our understanding of how best to harness its protective potential.

## Introduction

The use of anti-SARS-CoV-2 monoclonal antibodies for preexposure prophylaxis in kidney transplant recipients (KTRs) with a weak antibody response after vaccination has gained traction. Casirivimab–imdevimab combination (Ronapreve™, Roche Regeneron) has been shown to confer satisfactory protection against the delta variant.^1,2^ However, both Ronapreve™ and other antibodies have limited neutralizing activity against the current variants of concern (VOC) sublineages (omicron BA.1, BA.1.1 and BA.2). In contrast, cilgavimab–tixagevimab combination (Evusheld™, Astra Zeneca) retains a partial neutralizing activity against omicron *in vitro*^3–5^. Based on these data, health authorities have authorized the use of Evusheld™ for preexposure prophylaxis in immunocompromised patients with a weak anti-SARS-CoV-2 antibody response after vaccination. The level of clinical protection offered by this strategy remains ill determined as clinical trials on Evusheld™ were performed before the emergence of omicron.^6^ In this study, we report a case series of KTRs who developed omicron infection despite preexposure Evusheld™ administration.

## Methods

All procedures and visits occurred at the Strasbourg and Lyon University Hospitals (France). Intramuscular prophylactic injections of Evusheld™ (150 mg tixagevimab and 150 mg cilgavimab) were offered to KTRs as of December 28, 2021. The date of last follow-up was March 13, 2022. The diagnosis of COVID-19 was based on RT-PCR of nasopharyngeal swabs and genome sequencing was performed when suitable samples were available. The anti–receptor-binding domain (RBD) IgG response and neutralizing activity against the omicron BA.1 variant were assessed within the first 30 days after Evusheld™ injection and no later than the first seven days after the onset of COVID-19. All procedures complied with the Helsinki Declaration and were approved by the local Institutional Review Board (comité d’éthique, Université de Strasbourg, Strasbourg, France, reference number: CE-2021-9).

## Results

Of the 416 KTRs who received prophylactic injections of Evusheld™, 39 (9.4%) developed COVID-19 (Table 1). All had been previously vaccinated against SARS-CoV-2 with mRNA vaccine but failed to develop a protective humoral response. The median time elapsed from Evusheld™ injections to the onset of COVID-19 was 20 days (IQR: 9.5−34.5). With the exception of one patient, all KTRs were symptomatic. Hospitalization was required for 14 patients (35.9%) of whom three were transferred to intensive care unit. Two KTRs died of COVID-19-related acute respiratory distress syndrome. SARS-CoV-2 sequencing was carried out in 15 cases (BA.1, n = 5; BA.1.1, n = 9; BA.2, n=1). Viral neutralizing activity of the serum was negative in the 12 tested patients, suggesting that this prophylaxis strategy provides insufficient protection against this variant of concern of SARS-CoV-2. Five patients had anti-RBD IgG titers <3500 BAU/mL. In the remaining seven patients, preexisting Ronapreve™ administration did not allow interpreting anti-RBD IgG levels.

**Table 1.** General characteristics of kidney transplant recipients (n = 39) who developed COVID-19 after preexposure prophylaxis with tixagevimab and cilgavimab (Evusheld™)

| Patient # | Sex | Age range (y) | Time elapsed from KT (y) | Number of vaccine doses | Time elapsed from Evusheld™ injection (d) | Upper respiratory symptoms | Fever, headache, myalgia, chills | Lower respiratory symptoms | HA | ICU | Death | SARS-CoV-2 variant | IgG RBD (BAU/mL) | Neutralizing capacity against Omicron BA.1 |
| --- | --- | --- | --- | --- | --- | --- | --- | --- | --- | --- | --- | --- | --- | --- |
| 2 | M | 71-80 | 1.1 | 3 | 16 | No | Yes | Yes | Yes | Yes | Yes | BA1.1 |  |  |
| 30 | F | 71-80 | 1.1 | 3 | 28 | Yes | Yes | No | Yes | Yes | Yes | BA.1 | 5128* |  |
| 21 | M | 61-70 | 10.6 | 3 | 26 | Yes | No | No | Yes | Yes | No | BA.1 |  |  |
| 3 | F | 51-60 | 4.6 | 3 | 5 | Yes | No | No | Yes | No | No |  |  |  |
| 7 | M | 51-60 | 1.5 | 4 | 35 | No | Yes | Yes | Yes | No | No |  |  |  |
| 13 | M | 71-80 | 0.24 | 3 | 30 | No | Yes | No | Yes | No | No |  | 1775 | Negative |
| 18 | M | 71-80 | 4.4 | 3 | 42 | Yes | Yes | No | Yes | No | No | BA1.1 | 9442* | Negative |
| 22 | M | 61-70 | 2.6 | 2 | 12 | Yes | Yes | No | Yes | No | No | BA1.1 |  |  |
| 23 | M | 61-70 | 2 | 2 | 36 | No | Yes | Yes | Yes | No | No | BA.1 | 4241* | Negative |
| 24 | F | 71-80 | 8.7 | 3 | 22 | No | Yes | Yes | Yes | No | No | BA.1 | 3786* | Negative |
| 33 | M | 71-80 | 1.4 | 3 | 16 | Yes | Yes | No | Yes | No | No |  |  |  |
| 34 | F | 51-60 | 13.3 | 2 | 62 | Yes | Yes | No | Yes | No | No |  | 522 |  |
| 35 | M | 51-60 | 0.19 | 4 | 57 | Yes | No | Yes | Yes | No | No |  | 2771* | Negative |
| 36 | M | 71-80 | 7.4 | 3 | 32 | No | Yes | Yes | Yes | No | No | BA1.1 | 2785 |  |
| 4 | F | 21-30 | 1.6 | 3 | 10 | No | Yes | Yes | No | No | No | BA1.1 | 10932* | Negative |
| 1 | M | 41-50 | 0.1 | 2 | 5 | No | Yes | Yes | No | No | No | BA1.1 | 2458 | Negative |
| 5 | M | 51-60 | 4.8 | 3 | 18 | Yes | Yes | No | No | No | No |  |  |  |
| 6 | F | 71-80 | 12.9 | 3 | 5 | No | Yes | No | No | No | No | BA1.1 | 1790 | Negative |
| 8 | F | 31-40 | 18.2 | 4 | 9 | Yes | Yes | No | No | No | No |  |  |  |
| 10 | M | 51-60 | 3.17 | 3 | 37 | Yes | Yes | No | No | No | No |  | 6800* | Negative |
| 14 | M | 31-40 | 32.9 | 4 | 21 | Yes | Yes | No | No | No | No |  | 3420 | Negative |
| 15 | F | 71-80 | 1.1 | 3 | 9 | Yes | Yes | No | No | No | No |  |  |  |
| 16 | M | 51-60 | 4.3 | 3 | 32 | Yes | Yes | No | No | No | No | BA1.1 | 5182* | Negative |
| 20 | M | 41-50 | 2.3 | 2 | 4 | No | Yes | No | No | No | No | BA1.1 |  |  |
| 25 | M | 51-60 | 2.9 | 3 | 20 | No | Yes | No | No | No | No |  |  |  |
| 26 | F | 51-60 | 2.6 | 2 | 34 | Yes | Yes | No | No | No | No |  |  |  |
| 27 | M | 61-70 | 3.8 | 4 | 5 | Yes | Yes | No | No | No | No |  |  |  |
| 28 | F | 61-70 | 1.6 | 3 | 6 | Yes | Yes | No | No | No | No |  |  |  |
| 29 | F | 51-60 | 4.7 | 3 | 12 | Yes | Yes | No | No | No | No |  |  |  |
| 31 | M | 51-60 | 2.2 | 3 | 40 | No | Yes | No | No | No | No |  | 1581 |  |
| 32 | F | 71-80 | 1.5 | 3 | 1 | Yes | Yes | No | No | No | No |  | 3570* |  |
| 11 | F | 71-80 | 3.2 | 3 | 36 | Yes | No | No | No | No | No | BA.1 | 5686* | Negative |
| 12 | M | 61-70 | 1.1 | 3 | 12 | Yes | No | No | No | No | No |  |  |  |
| 9 | M | 21-30 | 7.3 | 3 | 12 | Yes | No | No | No | No | No |  |  |  |
| 19 | M | 51-60 | 4.6 | 4 | 12 | Yes | No | No | No | No | No |  |  |  |
| 37 | F | 51-60 | 1.5 | 3 | 46 | Yes | No | No | No | No | No | BA.2 | 5212* |  |
| 38 | F | 51-60 | 14 | 3 | 22 | Yes | No | No | No | No | No |  |  |  |
| 39 | M | 61-70 | 16.5 | 3 | 47 | Yes | No | No | No | No | No |  |  |  |
| 17 | F | 18-20 | 2.3 | 3 | 6 | No | No | No | No | No | No |  |  |  |

Abbreviations: KT, kidney transplantation; HA, hospital admission; ICU, intensive care unit; d, days, y, years; M, male; F, female.
\*Patients who received Ronapreve™ prior to Evusheld™ (uninterpretable anti-RBD IgG levels)
Orange background: hospitalized patients; yellow background: symptomatic patients managed out of hospital; white background: asymptomatic patient.

## Discussion

In this study, we describe the occurrence of severe omicron infections despite a prophylactic treatment with Evusheld™. Notably, two study participants died of COVID-19.

Previous studies have shown that the BA.1.1 subvariant is characterized by a higher *in vitro* resistance to Evusheld™ than the BA.1 variant^4,5^. The former genotype was dominant in our cohort, which could partly explain the disappointing level of protection observed in these patients. This problem is however unlikely the only explanation since we also observed that none of the sera collected after administration of Evusheld™ were able to neutralize the BA.1 variant in vitro. The latter results suggest that intramuscular injections of a combination of 150 mg tixagevimab and 150 mg cilgavimab might not be sufficient to reach protective levels of anti-RBD antibodies in the circulation.

Our clinical findings confirm recent FDA recommendations, derived from *in vitro* models, underlining the necessity to increase the dose of Evusheld™.^4^ Further pharmacokinetic studies are warranted to determine what is the optimal dose of Evusheld™ for primary prophylaxis against COVID-19. Waiting for these results, KTRs should be advised to maintain sanitary protection measures and undergo vaccine boosters.

## Data Availability

All data produced in the present study are available upon reasonable request to the authors

## Conflict of Interest Disclosures

Sophie Caillard and Olivier Thaunat received consultant fees from Astra Zeneca. No other disclosures were reported.

## Notes

### Funding Statement

This study did not receive any funding

### Author Declarations

All procedures complied with the Helsinki Declaration and were approved by the Institutional Review Board (comit&eacute d'&eacutethique, Universit&eacute de Strasbourg, Strasbourg, France, reference number: CE-2021-9).

